# An Application of Logistic Formula to Deaths by COVID-19 in Japan

**DOI:** 10.1101/2020.09.15.20195081

**Authors:** Takesi Saito

## Abstract

A logistic formula in biology is applied to analyze deaths by COVID-19 for both of the first and the second waves in Japan. We then discuss ends of both waves and their mortality ratios. The meaning of population *N* in an epidemic is discussed.

## 1 Introduction

The SIR model [1] in the theory of infection is powerful to analyze an epidemic about how it spreads and how it ends [2-8]. The SIR model is composed of three equations for *S, I* and *R*, where they are numbers for susceptibles, infectives and removed, respectively. Three equations can be solved numerically, if two parameters *α* and *c* are given, where *α* is the basic reproduction number and *c* the removed ratio.

In Sec. 2 we summarize approximate solutions of the SIR equations, based on the logistic formula in biology [9], which have been derived in a previous work [10]. These approximate solutions have simple forms, so that they are very useful to discuss an epidemic. In Sec. 3 these solutions are applied to fix the basic reproduction number *α* and the removed ratio *c*, especially from data of accumulated number of deaths by COVID-19 for the first wave in Japan. Here, our policy is to use only data of deaths, but not of cases. The mortality ratio for the first wave is also calculated. In addition we discuss the meaning of population *N* in an epidemic. In Sec. 4 same things are given for the second wave. The final section is devoted to concluding remarks. In Appendix data of deaths for the first and the second waves are summarized.

## 2 A logistic curve from the SIR model

Equations of the SIR model [1] are given by

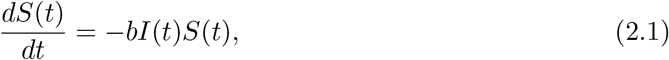

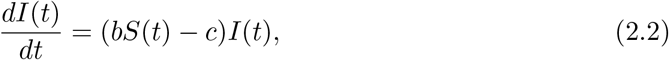

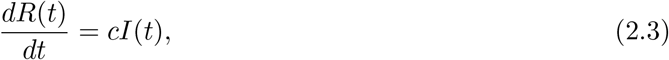

where *S, I* and *R* are numbers for susceptibles, infectives and removed, respectively,*b* the infection ratio and *c* the removed ratio. From Eqs. (2.1) and (2.3) we get *dS/dR* = −*αS*, (*α* = *b/c*), which is integrated to be

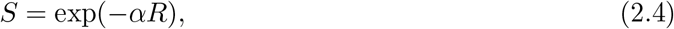

where *α* stands for the basic reproduction number. In the same way, from Eqs. (2.2) and (2.3), we have *dI/dR* = *αS* − 1 = *α* exp(−*αR*) − 1, which is integrated to be

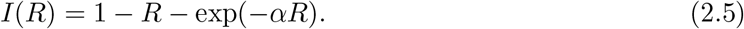

The solutions (2.4) and (2.5) satisfy boundary conditions *S* = 1 and *I* = 0 at *R* = 0. We normalize the total number to be unity, i.e., *S* + *I* + *R* = 1. Since *I*(*t*) is 0 when *t* → *∞*, we have 1 − *R*(*∞*) − exp[−*αR*(*∞*)] = 0 from Eq.(2.5). Hence, it follows a useful formula

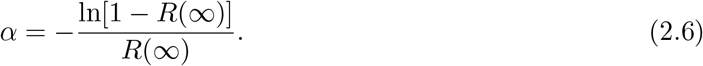

Some exact formulas at the infection peak *t* = *T* are summarized as follows:

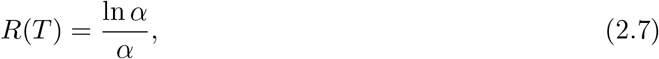

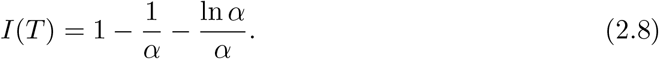

The first one is derived as follows: Since the peak point is given by *dI/dt* = *c*(*αS* − 1)*I* = 0 at *t* = *T*, we get *S*(*T*) = exp[−*αR*(*T*)] = 1*/α*. This proves Eq.(2.7). The second equation (2.8) directly follows by *I*(*T*) = 1 − *R*(*T*) − *S*(*T*) = 1 − ln *α/α* − 1*/α*.

Let us summarize logistic formulas for SIRs functions [10]. The third equation (2. 3) can be written as

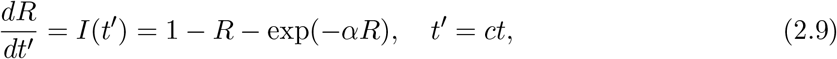

with *t* the true time. Let us expand the exponential factor in the second order of *x* = *αR*,

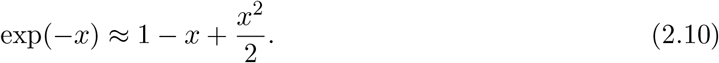

Then we have

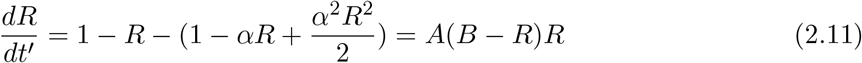

with *A* = *α* ^2^*/*2, *B* = 2(*α* − 1)*/α*^2^. This equation is a type of the logistic growth curve in the biology [9], easily solved as

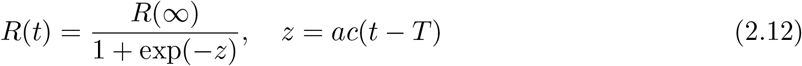

where *AB* = *α* − 1 = *a* and *B* = *R*(*∞*) = 2(*α* − 1)*/α*^2^. Inserting *R*(*t*) into *I*(*t*) = (1*/c*)*dR/dt*, we have

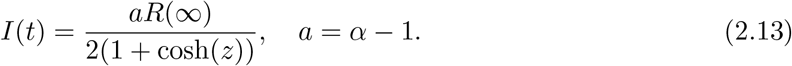

Note that peak values of *I*(*t*) and *R*(*t*) at *t* = *T* are given by

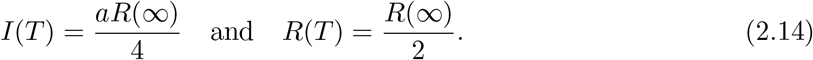

Hence we have a useful formula

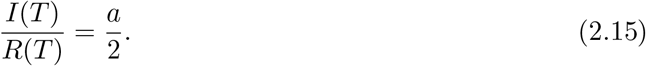

## 3 Application to COVID-19 for the first wave

We make use of data [11] of deaths rather than cases. It is our policy not to use data of cases. Define the mortality ratio *λ* by

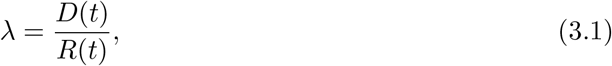

where *D*(*t*) is the accumulated number of deaths. For the population *N*, the total number of deaths at *t* is given by

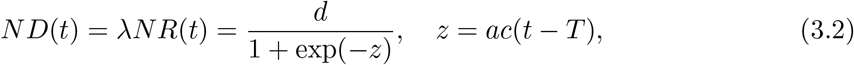

where *d* = *ND*(*∞*) = *λNR*(*∞*) stands for the final total number of deaths. Note that from Eq.(3.2) we have a theorem, *ND*(*T*) = *d/*2, which means the total number of deaths at the peak is just a half of the final ones.

Let us rewrite Eq.(3.2) into the form as

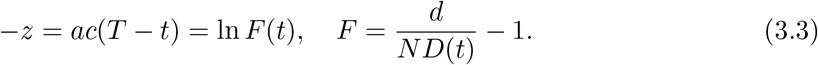

Accordingly, for different times *t*_1_, *t*_2_ and *t*_3_ we have

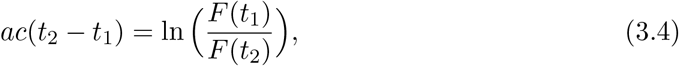

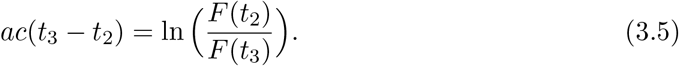

(*ND*(*t*) is the total number of deaths in the first wave at t in Japan, which is an average of 5-day deaths of *t* − 2, *t* − 1, *t, t* + 1, *t* + 2. The *t* is the number of date starting from Feb. 16) A use is made of data of deaths [11]. Substituting data on the Table 1 into Eqs.(3.4) and (3.5), and eliminating *ac*, we get

**Table 1:** Date and the total number of deaths in the first wave

| t | N D(t) |
| --- | --- |
| $t_1=95$(May 20) | 783 |
| $t_2=109$(June 3) | 904.8 |
| $t_3=123$(June 17) | 936 |

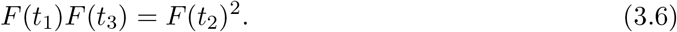

This is nothing but the equation for *d*, that is,

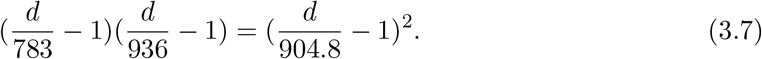

We find a solution of this equation to be d=984. According to the theorem *ND*(*T*) = *d/*2, the number of deaths at *t* = *T* is *d/*2 = 492. This means the peak day is May 2nd, as seen on data. The result of *d* = 984 is substituted into Eq.(3.4), then it follows

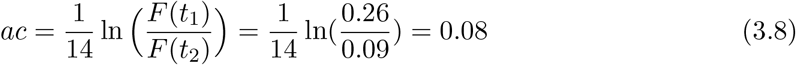

In the following we take *c* = 0.04, which is frequently quoted value. Then we have *a* = 0.08*/*0.04 = 2.00, that is, *α* = 1 + *a* = 3. Also, from the formula (2.6) it follows *R*(*∞*) = 0.94.

The original SIR equations are also solved by means of Excel. Their curves are drawn in Figure 1.

**Figure 1:**
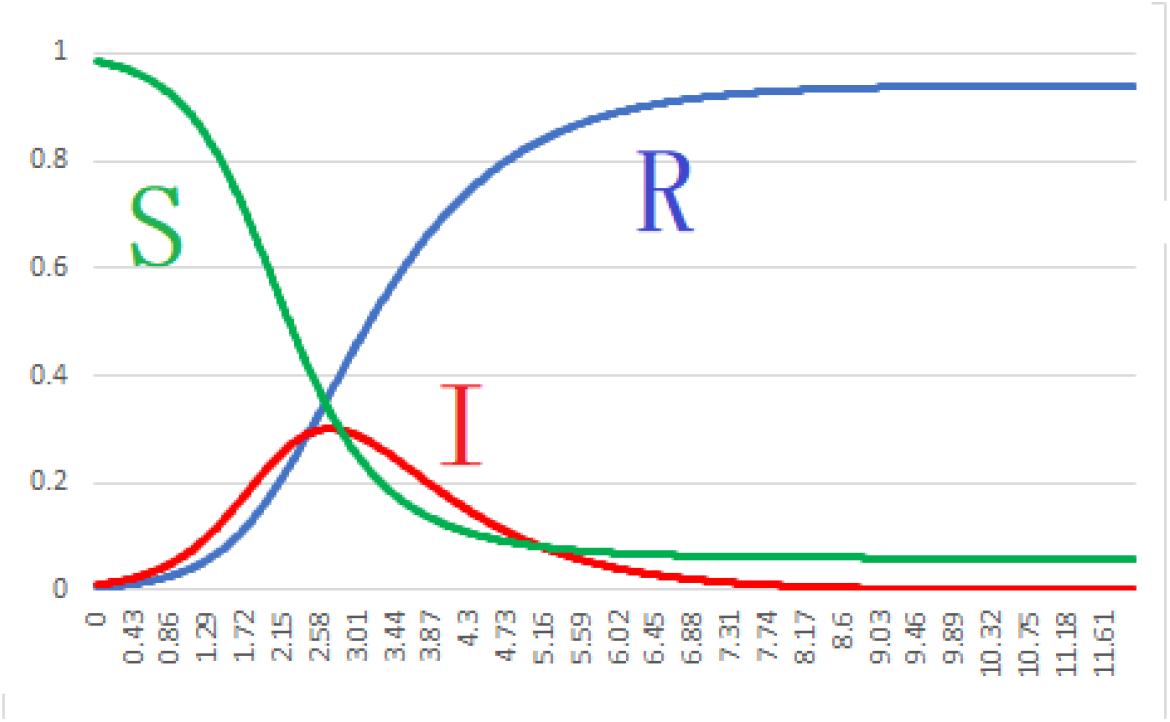
Graph of *S, I* and *R* for *α* = 3 and *c* = 0.04

From the curve of *I*(*t*′) we find that *t*′= 3.00 gives a peak, which is compared with the formula *t*′= *ct* = 0.04 *×* 77 = 3.08. This slightly differs from *t*′= 3.00. However, such a small difference may be allowed for a margin of error arising from our approximate method. After approximately 77 days later, the first epidemic will end around a middle of July(7/18) with the total number 984 of deaths.

Finally let us calculate the mortality ratio *λ* = *D*(*t*)*/R*(*t*). The removed number *R*(*t*) is given by *R*(*t*) = *r*(*t*) + *D*(*t*), where *r*(*t*) is the accumulated number of people discharged from hospital and *D*(*t*) is the accumulated number of deaths. We find its mean value is *< λ >*= 0.1 with *σ* = 0.002.

Here we have a comment about the population *N*. From Eq.(2.7) it follows *R*(*T*) = ln *α/α* = 0.37 for *α* = 3, whereas *NR*(*T*) = 4877 from the data. Hence we get *N* = 4877*/R*(*T*) = 4877*/*0.37 ≈ 13000. What is such a small population as *N* = 13000? In order to discuss this question, we divide the Japan population into two groups A and B, A is immune to the present epidemic and B is not immune the epidemic. We can add also people who completely quarantine themselves to A. We then conclude that such a small population as *N* = 13000 corresponds to B, because A is irrelevant to infection.

## 4 The second wave

The second wave of COVID-19 in Japan seems to start from around June 22. The virus is now called the Tokyo type.

(*ND*(*t*) is the total number of deaths with 3-day average in the second wave at *t*. The *t* is the number of date starting from July 20. We make use of data in the appendix, where the number of deaths in the second wave is completely separated from that of the first wave. It is shown that the first death of the second wave appears on July 20.)

The corresponding equation to Eq.(3.7) is, from Table 2,

**Table 2:** Date and the number of deaths in the second wave

| t | N D(t) |
| --- | --- |
| $t_1=32$(Aug. 20) | 171 |
| $t_2=38$(Aug. 26) | 239.3 |
| $t_3=38$(Sep. 1) | 309 |

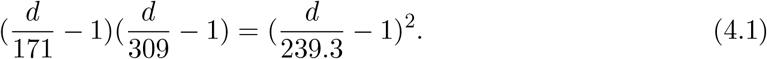

We have a solution *d* = 482, so that *d/*2 = 241 is the death number at the peak. Hence the peak day is Aug. 26 and *T* = 38, as seen from Appendix.

The result of *d* = 482 is substituted into Eq.(3.4), then it follows

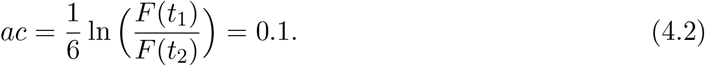

In the following we take *c* = 0.028.. Then we get *a* = 3.6, that is, *α* = 4.6. Once having the basic reproduction number *α* = 4.6 with the removed ratio *c* = 0.028, we can draw curves of *S, I* and *R* by means of Excel in Figure 2.

**Figure 2:**
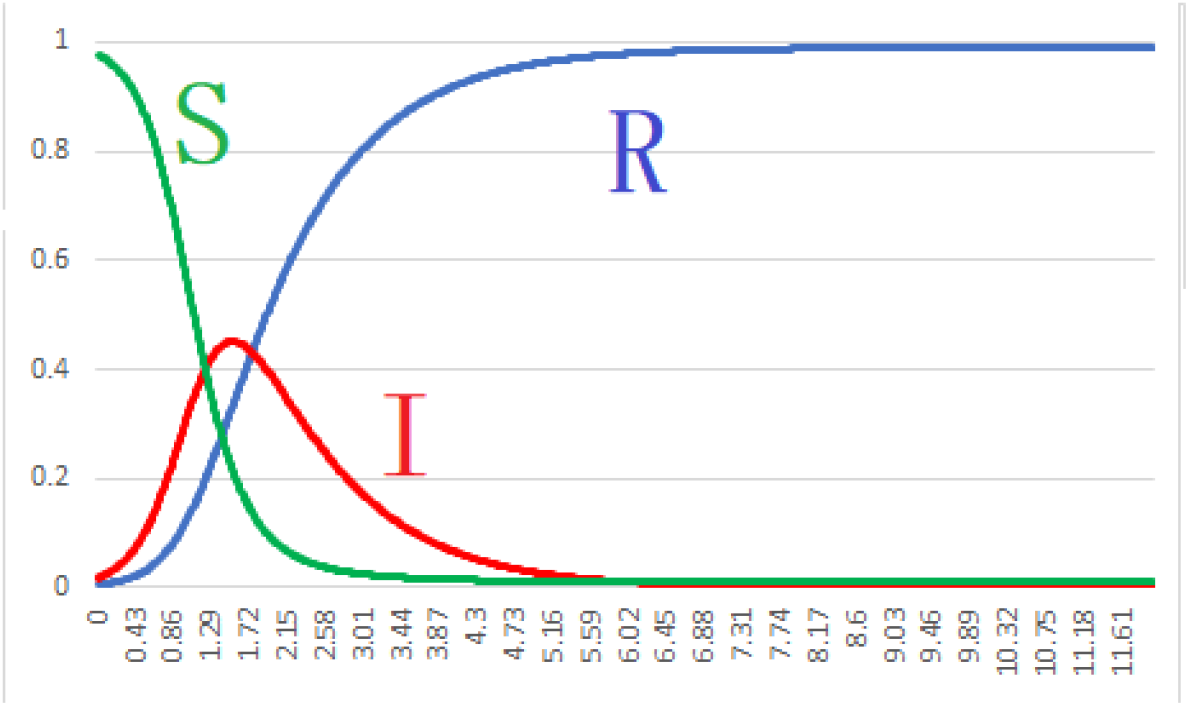
Graph of *S, I* and *R* for *α* = 4.6 and *c* = 0.028

From the curve of *I*(*t*′) we find that the peak day is *t*′= 1.51, which is compared with the formula *t* = *ct* = 0.028 *×* 38 = 1.06. This slightly differs from *t*′= 1.51. However, such a small difference may be allowed for a margin of error arising from our approximate method. Since the second infection begins from July 20, the *t*′= 1.51 corresponds with the true time *t* = 38 days, so that the peak day is Aug. 26, with the total number 241 of deaths. After approximately 38 days later, the epidemic will end around Oct. 3 with the total number 241 *×* 2 = 482 of deaths. One can see that a ratio of both infectives on peak days is *I*_2_*/I*_1_ = 3*/*2, where *I*_1_ and *I*_2_ stand for infectives of 1^st^ and 2^nd^ waves.

Finally let us calculate the mortality ratio *λ* = *D*(*t*)*/R*(*t*). We find that its mean value between July 20 and Aug. 22 is *< λ >*= 0.005 with *σ* = 0.0006.

## 5 Concluding remarks

Our logistic formulas have been applied to analyze deaths by COVID-19 in Japan. Our policy is to use only data of deaths, but not of cases.

For the first wave, the basic reproduction number *α* is determined to be *α* = 3 and the removed ratio *c* = 0.04. The corresponding curves of *S, I* and *R* are given in Figure 1. From the curve of *I*(*t*′) we find that the infection peak day is May 2^nd^, then after approximately 77 days later, the first epidemic will end around a middle of July(7/18) with the total number 984 of deaths. The mortality ratio *λ* = *D*(*t*)*/R*(*t*) is also calculated to be its mean value *< λ >*= 0.1 with *σ* = 0.002.

Here we have a comment about the population *N*. We find *N* being calculable from the formula *R*(*T*) = ln *α/α* = 0.37 for *α* = 3. Namely, from data we see *NR*(*T*) = 4877 to give *N* ≈ 13000. What is such a small population as *N* = 13000? In order to discuss this question, we divide the Japan population into two groups A and B, A is immune to the present epidemic and B is not immune the epidemic. We can add also people who completely quarantine themselves to A. We then conclude that such a small population as *N* = 13000 corresponds to B, because A is irrelevant to infection.

For the second wave, the basic reproduction number *α* is determined to be *α* = 4.6 and removed ratio *c* = 0.028. The corresponding curves of *S, I* and *R* are given in Figure 2. From the curve of *I*(*t*′) we find that the infection peak day is Aug. 26, then after approximately 38 days later, the second epidemic will end around Oct. 3, with the total number 482 of deaths. The mortality ratio *λ* is also calculated to be its mean value *< λ >*= 0.005 with *σ* = 0.0006. A ratio of both infectives of 1^st^ and 2^nd^ waves on peak days can be seen to be 2*/*3.

## Data Availability

I agree with the availability of all data referred.

## Acknowledgment

The author would like to express his deep gratitude to K. Shigemoto for many valuable discussions and big supports.

## Appendix

**Data of accumulated number of deaths in the second wave**

Appendix: Data of accumulated number of deaths in the second wave
| No. | month/day | $D_{12}$ | $D_2$ | $r_{12}$ | $r_2$ | $R_2$ | $\lambda_2$ |
| --- | --- | --- | --- | --- | --- | --- | --- |
| 0 | 7/19 | 984 | 0 | 19523 | 0 | - | - |
| 1 | 7/20 | 987 | 3 | 19921 | 398 | 401 | 75 |
| 2 | 7/21 | 988 | 4 | 20403 | 880 | 884 | 45 |
| 3 | 7/22 | 989 | 5 | 20787 | 1264 | 1269 | 40 |
| 4 | 7/23 | 991 | 7 | 21080 | 1557 | 1564 | 44 |
| 5 | 7/24 | 992 | 8 | 21319 | 1796 | 1804 | 44 |
| 6 | 7/25 | 995 | 11 | 21514 | 1991 | 2002 | 55 |
| 7 | 7/26 | 995 | 11 | 21868 | 2345 | 2356 | 47 |
| 8 | 7/27 | 997 | 13 | 22560 | 3037 | 3050 | 42 |
| 9 | 7/28 | 1000 | 16 | 23256 | 3733 | 3749 | 43 |
| 10 | 7/29 | 1003 | 19 | 23928 | 4405 | 4424 | 43 |
| 11 | 7/30 | 1005 | 21 | 24678 | 5155 | 5176 | 41 |
| 12 | 7/31 | 1010 | 26 | 25253 | 5730 | 5756 | 45 |
| 13 | 8/1 | 1010 | 26 | 25653 | 6130 | 6156 | 42 |
| 14 | 8/2 | 1011 | 27 | 26234 | 6711 | 6738 | 40 |
| 15 | 8/3 | 1015 | 31 | 26944 | 7421 | 7452 | 42 |
| 16 | 8/4 | 1021 | 37 | 27775 | 8252 | 8289 | 45 |
| 17 | 8/5 | 1025 | 41 | 28624 | 9101 | 9142 | 45 |
| 18 | 8/6 | 1032 | 48 | 29900 | 10377 | 10425 | 46 |
| 19 | 8/7 | 1038 | 54 | 31054 | 11531 | 11585 | 47 |
| 20 | 8/8 | 1039 | 55 | 32059 | 12591 | 12591 | 44 |
| 21 | 8/9 | 1046 | 62 | 32805 | 13282 | 13344 | 46 |
| 22 | 8/10 | 1051 | 67 | 33722 | 14199 | 14266 | 46 |
| 23 | 8/11 | 1058 | 74 | 34635 | 15112 | 15186 | 49 |
| 24 | 8/12 | 1062 | 78 | 35858 | 16335 | 16413 | 48 |
| 25 | 8/13 | 1072 | 88 | 37165 | 17642 | 17730 | 50 |
| 26 | 8/14 | 1084 | 100 | 38600 | 19077 | 19177 | 52 |
| 27 | 8/15 | 1087 | 103 | 39715 | 20192 | 20295 | 51 |
| 28 | 8/16 | 1098 | 114 | 40794 | 21271 | 21385 | 53 |
| 29 | 8/17 | 1114 | 130 | 41853 | 22330 | 22460 | 58 |
| 30 | 8/18 | 1127 | 143 | 43379 | 23856 | 23999 | 60 |
| 31 | 8/19 | 1143 | 159 | 44771 | 25248 | 25407 | 63 |
| 32 | 8/20 | 1154 | 170 | 45938 | 26415 | 26585 | 64 |
| 33 | 8/21 | 1168 | 184 | 47045 | 27522 | 27706 | 66 |
| 34 | 8/22 | 1175 | 191 | 47946 | 28423 | 28614 | 68 |
| 35 | 8/23 | 1180 | 196 | 48716 | 29193 | 29389 | 67 |
| 36 | 8/24 | 1195 | 211 | 49794 | 30271 | 30482 | 69 |
| 37 | 8/25 | 1208 | 224 | 51034 | 31511 | 31735 | 70 |
| 38 | 8/26 | 1225 | 241 | 52169 | 32646 | 32887 | 73 |
| 39 | 8/27 | 1237 | 253 | 53318 | 33795 | 34048 | 74 |
| 40 | 8/28 | 1254 | 270 | 54650 | 35127 | 35397 | 76 |
| 41 | 8/29 | 1263 | 279 | 55471 | 35948 | 36277 | 77 |
| 42 | 8/30 | 1278 | 294 | 56109 | 36586 | 36880 | 80 |
| 43 | 8/31 | 1295 | 311 | 57130 | 37607 | 37918 | 82 |
| 44 | 9/1 | 1306 | 322 | 57735 | 38212 | 38534 | 84 |
| 45 | 9/2 | 1318 | 334 | 58827 | 39304 | 39638 | 84 |

$D_{12}$ : Accumulated number of deaths for 1<sup>st</sup>+2<sup>nd</sup> waves
$D_2$ : Accumulated number of deaths for 2<sup>nd</sup> wave
$r_{12}$ : Accumulated number of discharged for 1<sup>st</sup>+2<sup>nd</sup> waves
$r_2$ : Accumulated number of discharged for 2<sup>nd</sup> waves
$R_2 = D_2 + r_2$ : Removed number for 2<sup>nd</sup> wave
$\lambda_2 = D_2/R_2$ : Mortality rate for 2<sup>nd</sup> wave( $\times 10^{-4}$ )

